# The influence of socioeconomic deprivation on dementia mortality, age at death and quality of diagnosis: a nationwide death records study in England and Wales 2001-2017

**DOI:** 10.1101/2020.09.28.20203000

**Authors:** Mark Jitlal, Guru NK Amirthalingam, Tasvee Karania, Eve Parry, Aidan Neligan, Ruth Dobson, Alastair J Noyce, Charles R Marshall

**Affiliations:** Preventive Neurology Unit, Wolfson Institute of Preventive Medicine, Queen Mary University of London, London UK; Department of Neurology, Barts Health NHS Trust, London UK; Department of Clinical and Movement Neurosciences, UCL Queen Square Institute of Neurology, London UK; Department of Neurology, Homerton University Hospital NHS Foundation Trust, London, UK; Department of Experimental & Clinical Epilepsy, UCL Queen Square Institute of Neurology, London, UK; Dementia Research Centre, UCL Queen Square Institute of Neurology, London, UK

**Keywords:** Alzheimer’s, dementia, deprivation, socioeconomic status, mortality, age at death, diagnosis

## Abstract

**Background:** Socioeconomic deprivation is postulated to be an important determinant of dementia risk, mortality, and access to diagnostic services. Nevertheless, premature mortality from other causes and under-representation of deprived individuals in research cohorts may lead to this effect being overlooked.

**Methods:** We obtained Office of National Statistics (ONS) mortality data where dementia was recorded as a cause of death in England and Wales from 2001 to 2017, stratified by age, diagnosis code and UK Index of Multiple Deprivation (IMD) decile. We calculated standardised mortality ratios (SMR) for each IMD decile, adjusting for surviving population size in each IMD decile and age stratum. In those who died of dementia, we used ordinal logistic regression to examine the effect of deprivation on likelihood of being older at death. We used logistic regression to test the effect of deprivation on likelihood of receiving a diagnosis of unspecified dementia, a proxy for poor access to specialist diagnostic care.

**Results:** 578,623 deaths due to dementia in people over the age of 65 were identified between 2001-2017. SMRs were similar across the three most deprived deciles (1-3) but progressively declined through deciles 4-10 (Mean SMR [95%CI] in decile 1: 0.528 [0.506 to 0.550], decile 10: 0.369 [0.338 to 0.400]). This effect increased over time with improving ascertainment of dementia. In 2017, 14,837 excess dementia deaths were attributable to deprivation (21.5% of the total dementia deaths that year). There were dose-response effects of deprivation on likelihood of being older at death with dementia (odds ratio [95%CI] for decile 10 (least deprived): 1.31 [1.28 to 1.33] relative to decile 1), and on likelihood of receiving a diagnosis of unspecified dementia (odds ratio [95%CI] for decile 10: 0.78 [0.76 to 0.80] relative to decile 1).

**Conclusions:** Socioeconomic deprivation in England and Wales is associated with increased dementia mortality, younger age at death with dementia, and poorer access to specialist diagnosis. Reducing social inequality may be an important strategy for prevention of dementia mortality.

## Introduction

Persistent and widening socioeconomic inequality in the United Kingdom is associated with negative health outcomes including excess premature mortality in those who are more deprived^(1)^. However, the effect of this on dementia mortality across the United Kingdom has not been systematically examined.

Socioeconomic deprivation has previously been shown to be a risk factor for dementia^(2-6)^. Various factors have been hypothesised to mediate this relationship, including cognitive reserve, education, diet, vascular risk factors, stress and access to healthcare^(7)^. Deprivation is closely linked to education, which has been more widely studied as a risk factor for dementia, but some evidence suggests that wealth and area-based indices of deprivation may be more important than education when all are taken into account^(8, 9)^. Deprivation has also been associated with earlier death from dementia and with reduced access to good dementia care^(10, 11)^.

There are obstacles to examining the effect of deprivation on dementia outcomes, and its importance as a risk factor is therefore often overlooked^(8)^. Cohort studies tend to under-represent more deprived participants, while in population studies survival bias and incomplete ascertainment of cases in more deprived groups due to healthcare inequalities may lead to underestimation of the influence of deprivation^(12-16)^.

We used nationwide death certificate data from all of England and Wales during a 17-year period from 2001 to 2017. Deprivation was measured using the Index of Multiple Deprivation (IMD). We were able to partly mitigate the influence of premature mortality from other causes by adjusting for surviving population size within each IMD decile in each year. The primary aim of the study was to test the hypothesis that age-standardised mortality from dementia would be higher in more deprived deciles, and that this effect would become greater over time due to disproportionate improvements in ascertainment in the more deprived deciles. Furthermore, we hypothesised that those dying of dementia would be younger on average in more deprived deciles. Finally, we hypothesised that more deprived deciles would be more likely to have an unspecified dementia diagnosis (as opposed to any specified dementia syndrome), reflecting poorer access to specialist diagnostic services^(17)^.

## Methods

### Data Selection

Mortality data were obtained from the Office for National Statistics (ONS). A search of dementia deaths was conducted to obtain data between 2001-2017 (from the start of ICD-10 coding to the latest year available at the time of data access), for those aged 65 and over in England and Wales where dementia was listed as a cause of death according to ONS coding algorithms. The lower age limit of 65 was chosen because this is the standard age used to dichotomise early and late onset dementia. Below this age, dementia is much more likely to be genetic in aetiology, less likely to be influenced by life course factors, and the numbers of deaths are very small resulting in imprecise mortality ratio estimates. The number of deaths were split by age group (65-69; 70-74; 75-79; 80-84; 85-89; 90+), Index of Multiple Deprivation (IMD) decile (1: most deprived), year and specific dementia type at death (Alzheimer’s disease; Unspecified dementia; Vascular dementia; Other specified dementia), according to the ICD-10 code. A full list of ICD-10 codes and their diagnostic groupings is available in Supplementary Table 1. The population size for each age band and IMD decile, per year, was also obtained from the ONS.

### Defining deprivation

Deprivation was measured using the UK Index of Multiple Deprivation (IMD). This is an area-based measure of socioeconomic status that ranks every lower-layer super output area (LSOA; a small geographical area with on average 1500 inhabitants) in England and Wales, taking into account seven domains of deprivation: income, employment, education, health, crime, barriers to housing and services, and living environment. Each LSOA is then assigned to a national decile of deprivation.

### Statistical Methods

#### Effect of deprivation on dementia mortality

Population size was aggregated across age bands and the number of deaths across age bands and dementia type. Mortality rates were calculated for each IMD-year level. IMD decile 1 in 2017 was used as the reference category and the standardised mortality ratio (SMR) was calculated for all levels. As we wished to model the effect of IMD on SMR over time, linear models are not appropriate for this data, given the non-linear relationship between time and SMR for each IMD decile. Instead we used a generalised additive model (GAM), which uses smoothing functions to model non-linear relationships. In our case we modelled year as a smooth term. We calculated excess deaths attributable to deprivation in 2017 by determining the expected deaths in each IMD decile based on the SMR for decile 10, and then subtracting this from the observed number of deaths.

### Effect of deprivation on age at death

In order to examine the effect of IMD on age at death from dementia, the number of deaths due to dementia was aggregated across year and dementia type. We used an ordinal logistic regression model to determine the effect of IMD on age at death with dementia, expressed as the cumulative odds of being in any older age group at death. For each decile, the percentage of deaths from that decile that occurred in each age-group was also calculated.

### Effect of deprivation on dementia diagnosis

In order to examine the effect of IMD on whether the subtype of dementia at death was specified or not (a proxy for access to appropriate specialist care^(17)^, the number of deaths was aggregated across age and year, and categorised as unspecified dementia or any specified dementia. We used a logistic regression model to determine the effect of IMD on the odds of having a diagnosis of unspecified dementia versus any specified dementia diagnosis.

Analyses were conducted using R (version 3.6.2).

### Data availability statement

All data are publicly available from the Office of National Statistics: https://www.ons.gov.uk/

## Results

There were 578,623 recorded deaths due to dementia from 2001-2017 in England and Wales, with an overall SMR of 3.69 per 1000 people over the age of 65. 351,438 (61%) of dementia deaths were recorded as unspecified dementia, whereas only 137,477 (24%) deaths were recorded as being due to Alzheimer’s disease. The total number of deaths recorded by year, IMD decile, dementia type and age-group are available in Supplementary Table 2.

### Effect of deprivation on mortality

In general, the SMR increased over time, with higher standardised mortality in more deprived deciles (Figure 1). There was a marked increase in mortality between 2010 and 2011 (median increase in SMR=0.19, range 0.16 to 0.24) and a smaller increase seen between 2014 and 2015 (median increase in SMR=0.12, range 0.09 to 0.16). These coincided with ONS recoding of vascular dementia (2010-2011)^(18)^ and incentivisation of dementia diagnosis recording in primary care (2014-15)^(19)^. A GAM was implemented to assess the trend of SMR across deprivation, by smoothing the non-linear relationship seen between year and SMR (Table 1a). This showed that IMD deciles 4-10 have a reduced SMR compared to IMD decile 1, after accounting for year. There was stronger statistical evidence of a difference in mortality as the deciles increased, with the least deprived having the lowest mean SMR (0.369, 95% CI: 0.338 to 0.400); a decrease of 0.159 from IMD1; p < 0.001). In 2017 there were 14,837 excess dementia deaths attributable to deprivation (21.5% of all recorded dementia deaths that year).

**Figure 1:**
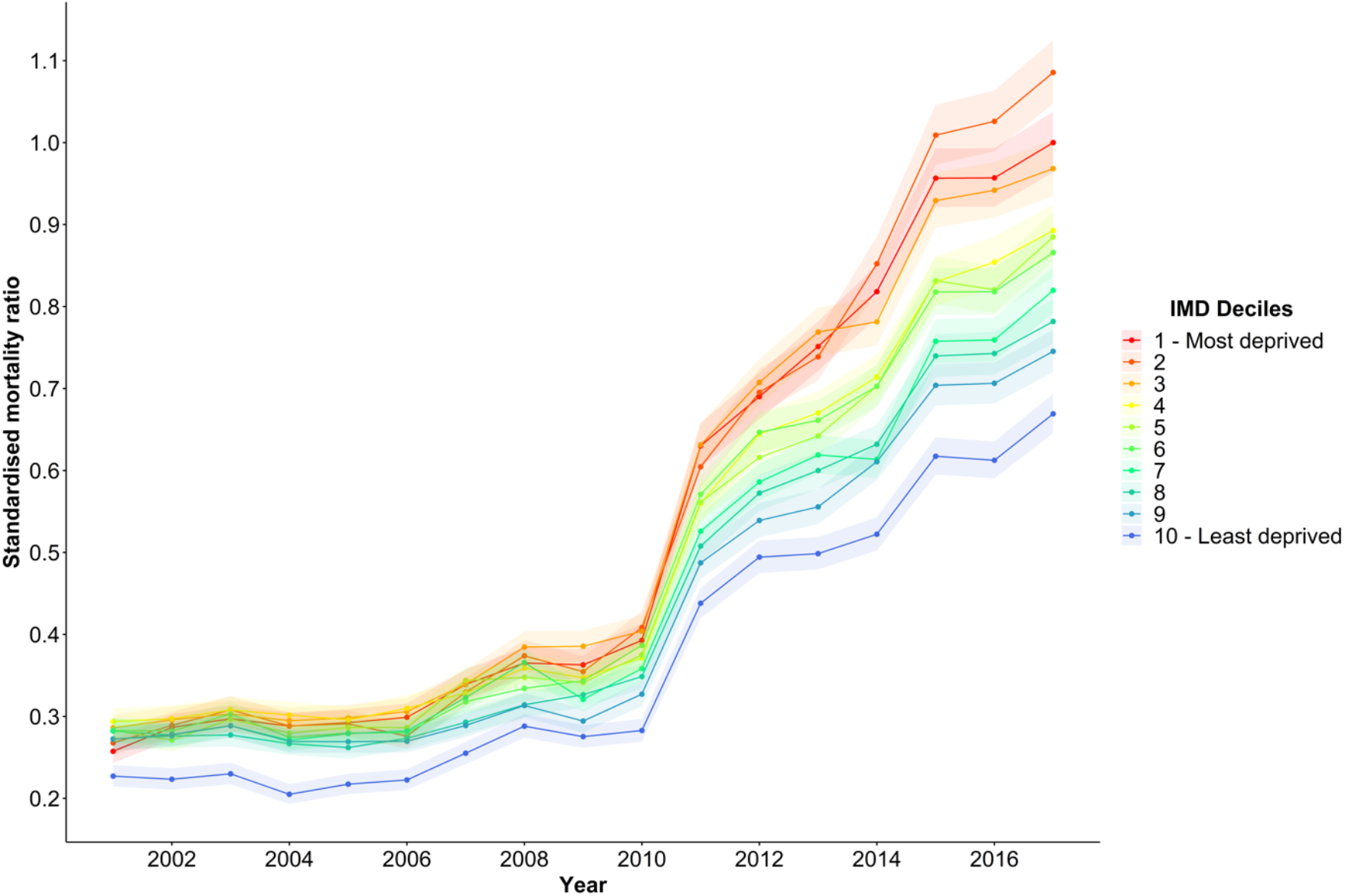
Standardised Mortality Ratios for IMD deciles over time. The figure shows SMRs for each IMD decile for population aged 65 or above in England and Wales, with IMD decile 1 in 2017 as the reference category. Shaded areas represent 95% confidence intervals for SMR estimates. IMD, UK Index of Multiple Deprivation.

### Effect of deprivation on age at death

An ordinal logistic regression analysis was implemented to investigate the influence of IMD on the age at death with dementia (Table 1b). This indicated that as deprivation decreases, then the odds of dying at an older age increase. For example, the least deprived decile had the greatest OR of 1.31 (95% CI: 1.28 to 1.33), indicating that the odds of dying at an older age is 31% greater than for the most deprived decile.

Amongst each IMD decile the proportion of patients dying in each age category was examined (Figure 2). This showed that a greater proportion of the most deprived deciles died in the younger age groups (e.g. 1.6% of deaths in the most deprived decile occurred in the 65-69 age group, compared to 1.3% of the least deprived [p<0.001]). This relationship persisted up until the 80-84 age group. A higher proportion of dementia deaths in the least deprived deciles occurred over the age of 90 (38.8% of the least deprived decile compared to 33.1% of the most deprived decile).

**Figure 2:**
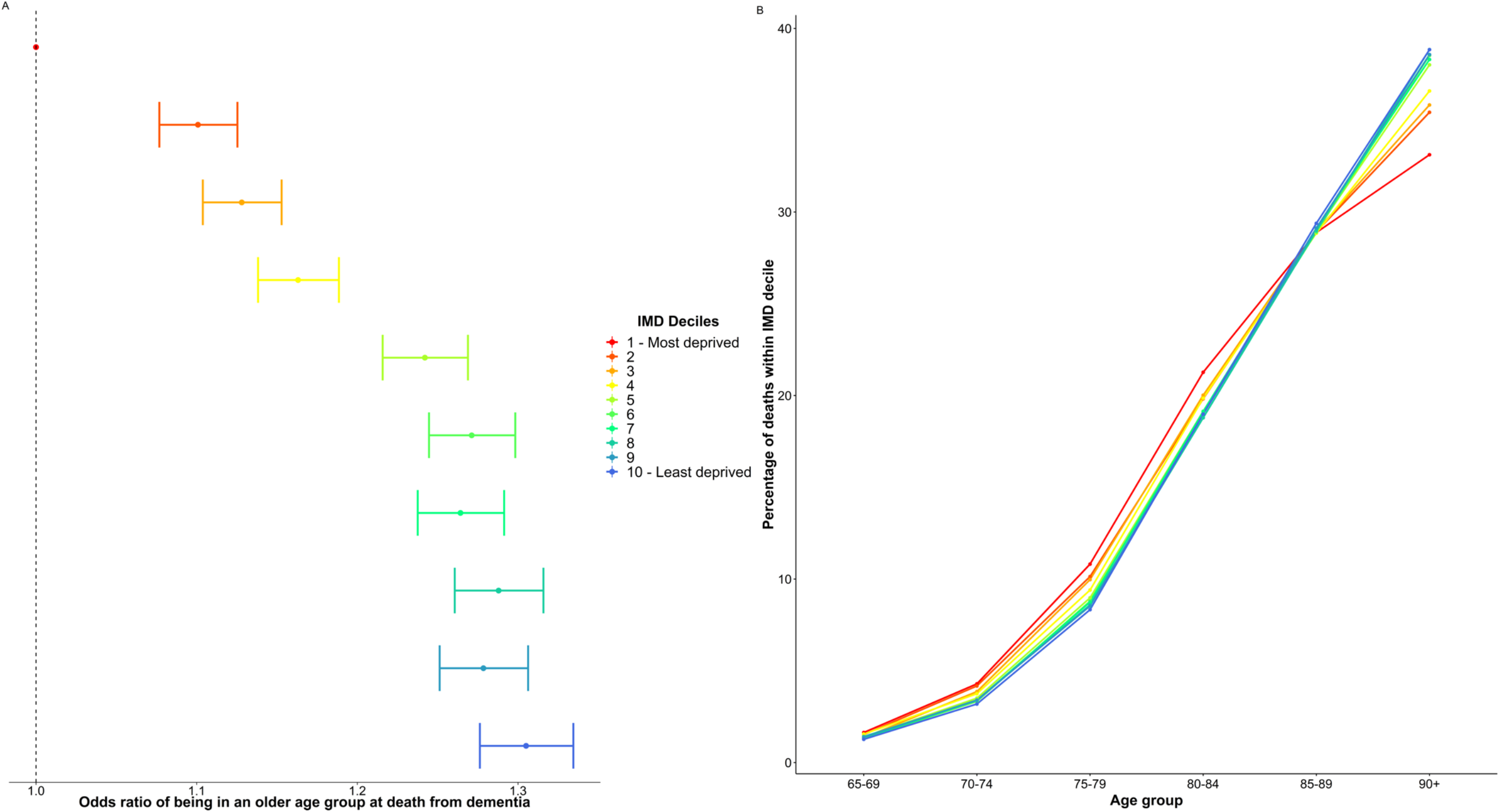
The effect of deprivation on age at death with dementia. A: Odds ratios (95% CI) of dying in an older age group according to IMD decile among those who died of dementia aged over 65 in England and Wales between 2001 and 2017. B: Percentage of dementia deaths within each IMD decile occurring in each age group in England and Wales 2001-2017 IMD, UK Index of Multiple Deprivation.

### Effect of deprivation on dementia diagnosis

Decreasing deprivation was associated with a dose-dependent decrease in the odds of having an unspecified dementia diagnosis compared to any specified aetiology (Table 1c). For example, people from the most affluent areas had 22% lower odds of having an unspecified diagnosis at death (95% CI: 20 to 24%), compared to those in the poorest areas.

## Discussion

In this study, using routinely collected death certificate diagnoses in England and Wales between 2001 to 2017, we demonstrate that greater socioeconomic deprivation is associated with higher dementia mortality and this effect appears to be increasing over time. These findings add to mounting evidence that socioeconomic status is an important determinant of dementia risk^(2-6)^. Our results highlight that deprivation should be considered a major target in public health approaches aimed at reducing the population burden of dementia.

The steadily rising dementia SMR evident in these data despite falling age-specific dementia incidence during the same time period is likely to be due to improving ascertainment^(16)^. Particularly large year-on-year increases in SMR coincided with known improvements in ascertainment related to recoding of vascular dementia by ONS (2011) and incentivisation of dementia diagnosis recording in primary care (2015)^(18, 19)^.

We also found that disparities in dementia mortality according to deprivation increased steadily over this time period. This could be because of disproportionately poor ascertainment in more deprived populations in earlier years, as a result of which improving ascertainment has begun to reveal the true scale of the effect. Of greater public health concern would be the alternative explanation that persistent and widening inequality in England and Wales is having an increasingly deleterious effect on brain health.

Being more deprived was associated with younger age at death in those dying from dementia. This supports the view that the excess in premature mortality found previously in UK death certificate data is partly due to dementia deaths^(1)^. It also supports previous findings that lower socioeconomic status is associated with diagnosis of dementia at a younger age and earlier mortality from dementia^(10, 20)^.

The finding that more deprived deciles were more likely to receive a diagnosis of unspecified dementia implies that these groups had poorer access to specialist diagnostic services^(17)^.

This is consistent with previous evidence showing that more deprived patients access services later, and are less likely to be prescribed anti-dementia drugs, implying that they are receiving lower quality care^(11, 21)^. These findings raise important challenges for the design of memory clinic services in order to ensure equitable access, diagnosis and treatment. From a clinical perspective, it is likely that poorer quality of diagnosis in more deprived patients means that they are being disadvantaged in terms of prognostication, counselling, planning of future care, access to appropriate symptomatic treatments and opportunities to participate in research. The under-representation of participants of low socioeconomic status is a key challenge for the validity and generalisability of dementia research including clinical trials, and ensuring more timely and accurate diagnosis would be an important first step to mitigate this.

The strengths of this work are that it is a large nationwide study that is perfectly representative of the population. Moreover, the ability to adjust for the surviving population size in each deprivation decile and age group made it possible to mitigate against survival effects to some extent. This is evident when comparing the trends for numbers of dementia deaths in each IMD decile (which are highest in decile 6, and similar in deciles 1 and 10; see Supplementary Table 2) to the trend of SMR after adjusting for surviving population size, which is highest in more deprived deciles. These aspects are likely to explain why we found important effects of deprivation on mortality and age at death when these were not previously detected in a UK population study over a similar time period^(13)^.

There are important limitations to the current study. Chief among these is the ascertainment of dementia through coding in death certificates. This is likely to have high positive predictive value for all-cause dementia, but lower positive predictive value for specific dementia subtypes, and is likely to lead to incomplete ascertainment of dementia cases^(22)^. It is noteworthy, however, that improved ascertainment over time appears to have enhanced the size of the effect, suggesting that incomplete ascertainment would tend to bias towards the null in this study. Although we were able to adjust for surviving population size, this does not completely correct for survival bias. In particular, those with premature mortality from other causes are likely to have comorbidities that increase dementia risk, and would therefore be more likely to develop dementia in later years than the background population. It is possible therefore that even in this very comprehensive dataset, the effect of deprivation is underestimated. Another major limitation is the use of an area-based summary measure of deprivation. This does not allow any inferences to be made about which aspects of deprivation are mediating the effects. In particular, it is possible that deprivation here is simply serving as a proxy for lower educational attainment, which is known to be an important determinant of dementia risk^(23)^. However, recent evidence suggests that the reverse may be true, i.e. that education acts as a proxy for deprivation more generally in studies on dementia risk ^(9, 24, 25)^. Moreover, the evidence for an influence of low education principally relates to having no secondary level education or being illiterate^8, 26)^. Secondary education has been compulsory in England and Wales since the Fisher Act of 1918 and literacy rates in the United Kingdom are around 99%, hence it is unlikely that the large effects seen here could be attributable to a very small proportion of the population who were illiterate or had no secondary education. Finally, with these data, we are unable to account for changing socioeconomic status over time, or for internal migration, and therefore we are unable to assess when in the life course deprivation is mediating the observed effects. This may be important, as previous work has suggested both cumulative effects across the life course and differential effects at different ages, with cognitive decline most strongly linked to wealth in later life^27, 28)^.

Future work should focus on addressing the issues of under-representation and survival bias in well-phenotyped cohorts that allow for more detailed analysis of the factors mediating the influence of deprivation on risk of dementia. Important outstanding questions include clarifying which aspects of deprivation are having the most important effects, when in the life course these effects are occurring, and how the effects are mediated. Whereas some studies have suggested that cognitive reserve is the most important mediator of increased dementia risk^(29)^, others have suggested that the effect is mediated by other modifiable risk factors, especially vascular risk^(24)^. Other suggested links between socioeconomic status and dementia outcomes such as stress, diet and air pollution have not been comprehensively evaluated as mediators^(7)^. A more complete understanding of these issues will allow for the design of public health interventions that target the most deleterious components of deprivation at the appropriate time in life with a view to preventing future dementia.

## Conclusions

Socioeconomic deprivation in England and Wales is associated with higher mortality from dementia, younger age at death with dementia and poorer quality of diagnosis. This suggests that political failures to combat persistent and widening socioeconomic inequality in the UK are contributing to the rising tide of dementia. In the context of enormous and growing societal costs of dementia, and the failure of disease modifying therapies, there should be added impetus to address deprivation with a view to promoting lifelong brain health and potentially preventing dementia.

## Data Availability

All data are publicly available from the Office of National Statistics: https://www.ons.gov.uk/

## Author Contributions

The corresponding author is responsible for the overall content as guarantor, and attests that all listed authors meet authorship criteria and that no others meeting the criteria have been omitted.

## Competing Interests

All authors have completed the ICMJE uniform disclosure form at www.icmje.org/coi_disclosure.pdf and declare: all authors had financial support from Bart’s Charity for the submitted work; no financial relationships with any organisations that might have an interest in the submitted work in the previous three years; no other relationships or activities that could appear to have influenced the submitted work.

## Table

**Table 1.** The effect of deprivation on dementia mortality, age at death and quality of diagnosis. The table shows the effects of IMD decile on a) standardised mortality ratio for dementia (averaged across years 2001-2017), b) cumulative odds of dying in an older age group and c) odds of receiving a diagnosis of unspecified dementia in England and Wales 2001-2017. CI, confidence interval; IMD, UK Index of Multiple Deprivation; OR, odds ratio; SMR, standardised mortality ratio

|  | a) | b) | c) |
| --- | --- | --- | --- |
| IMD decile: | Mean SMR (95% CI) | OR of being in an older age group at death (95% CI) | OR of 'Unspecified dementia' versus any specified (95% CI) |
| Most deprived - 1 | 0.528 (0.506, 0.550) | 1 | 1 |
| 2 | 0.541 (0.510, 0.572) | 1.10 (1.08, 1.13) | 0.98 (0.95, 1.00) |
| 3 | 0.531 (0.500, 0.562) | 1.13 (1.10, 1.15) | 0.97 (0.95, 1.00) |
| 4 | 0.493 (0.462, 0.524) | 1.16 (1.14, 1.19) | 0.96 (0.94, 0.99) |
| 5 | 0.481 (0.450, 0.512) | 1.24 (1.22, 1.27) | 0.91 (0.89, 0.94) |
| 6 | 0.480 (0.450, 0.512) | 1.27 (1.24, 1.30) | 0.87 (0.85, 0.89) |
| 7 | 0.455 (0.424, 0.486) | 1.26 (1.24, 1.29) | 0.83 (0.81, 0.85) |
| 8 | 0.440 (0.409, 0.471) | 1.29 (1.26, 1.32) | 0.82 (0.80, 0.84) |
| 9 | 0.425 (0.394, 0.456) | 1.28 (1.25, 1.31) | 0.81 (0.79, 0.83) |
| Least deprived - 10 | 0.369 (0.338, 0.400) | 1.31 (1.28, 1.33) | 0.78 (0.76, 0.80) |

## Notes

### Competing Interest Statement

The authors have declared no competing interest.

### Funding Statement

The Preventive Neurology Unit is supported by a grant from Bart's Charity

### Author Declarations

Ethical approval not required

